# Relationship between adherence to the 2019 Canada’s Food Guide recommendations on healthy food choices and nutrient intakes in older adults

**DOI:** 10.1101/2023.02.13.23285868

**Authors:** Didier Brassard, Stéphanie Chevalier

**Affiliations:** School of Human Nutrition, McGill University, Québec, Canada; Research Institute of the McGill University Health Centre, McGill University, Québec, Canada

**Keywords:** older adults, 24-hour dietary recalls, CFG, HEFI-2019, Canada’s Food Guide, dietary guidelines, healthy eating food index, Canadian Community Health Survey, CCHS.

## Abstract

**Background:** Following Canada’s food guide (CFG) recommendations should ensure adequate nutrient intakes. Older adults have increased needs for certain nutrients and nutrient density; the extent to which adherence to CFG recommendations can help reduce inadequate nutrient intakes is unknown.

**Objective:** Our aim was to assess the relationship between adherence to CFG recommendations on healthy food choices and intake of key nutrients in adults 65 years and older from the Canadian Community Health Survey (CCHS) 2015 - Nutrition.

**Methods:** Secondary analysis of data from 4,093 older adults of the CCHS 2015 - Nutrition (mean age, 73.6 years, 54% females). Dietary intakes were measured using an interviewer-administered 24-hour dietary recall including one repeat in a subsample (42%). The National Cancer Institute multivariate method was used to estimate usual (i.e., long-term) dietary intakes. Adherence to CFG recommendations was measured using the Healthy Eating Food Index (HEFI)-2019 score. Simple linear and logistic regression models estimated the effect of increased HEFI-2019 score on usual nutrient intakes and the prevalence of inadequate nutrient intakes (i.e., below the estimated average requirements), respectively.

**Results:** Compared with the prevalence of inadequate intakes at median HEFI-2019 score (46.4/80 points), a higher HEFI-2019 (+11 points) was associated with reductions in the prevalence of inadequate intakes of magnesium, vitamin B6, and protein (-19.8% [95%CI: -30.8, -8.9], -12.7% [95%CI: -22.5, -3.0], and -4.7% [95%CI: -9.4, -0.1], respectively). In contrast, data for higher HEFI-2019 scores were compatible with increased prevalence of inadequate intakes of folate, vitamin D, and calcium (4.0% [95%CI: -8.4, 16.3], 2.6% [95%CI: 1.1, 4.0], and 2.3% [95%CI: -3.0, 7.5], respectively).

**Conclusions:** Based on dietary intakes of Canadian older adults in 2015, increasing the degree of adherence to CFG recommendations on healthy food choices may reduce nutrient intake inadequacy for most key nutrients except folate, vitamin D and calcium.

## Introduction

Previous editions of the Canada’s Food Guide (CFG) informed on food choices to prevent malnutrition as the primary goal. Recommendations of the CFG-2007 were expressed in terms of number of servings to consume every day for the broad categories of foods: Vegetables and fruits, Grains, Dairy and Alternatives, and Meat and Alternatives (1, 2). The CFG-2007 recommended daily servings based on comprehensive diet simulations, thus ensuring that Dietary Reference Intakes (DRI) were met (2). The latest CFG (2019) adopted a different approach and primarily aims at chronic disease risk reduction (3–6). In addition, recommendations in CFG-2019 (e.g., “eat more often …”) are more flexible than those of CFG-2007 and rely on proportions of food categories to constitute a healthy plate, without recommended quantitative daily servings.

The release of the CFG-2019 elicited positive reactions (7, 8), but also raised concerns on nutrient adequacy. For example, preliminary modeling of eating patterns consistent with the CFG-2019 plate snapshot revealed that adherence may be insufficient to meet calcium and vitamin D requirements (9). The ability of consumers and policymakers to properly implement CFG-2019 recommendations on protein foods was also found unclear (10). In the Canadian Community Health Survey (CCHS) 2015 – Nutrition, animal-based protein foods contributed more than two thirds of total protein intake (11) as well as to overall nutrient intakes (12), which contrasts with CFG-2019 recommendations aiming at increasing intake of plant-based foods. Accordingly, following CFG-2019 recommendations while failing to include nutrient-dense foods and beverages could amplify the proportion of the population with inadequate nutrient intakes, especially for certain age groups at higher risk. In that regard, maintaining adequate nutrient intakes is a challenge for older adults (13, 14). Older adults face social (e.g., loneliness, inability to buy or prepare foods) and physiological (e.g., changes in taste, loss of appetite, malabsorption) barriers to consuming a nutrient-dense diet (14). Notably, calcium, vitamin D, fibre, potassium and protein were identified as nutrients of concern among older adults in the United States 2015 Dietary Guidelines report (15). Similarly, a high proportion of long-term care residents in Canada had inadequate calcium, folate, vitamin B6 and magnesium intakes (16). In sum, the one-size-fits-all recommendations in CFG-2019 may not be well-suited for older adults who face unique challenges to healthy eating compared with other strata of the population.

Whether adherence to CFG-2019 recommendations on healthy food choices fulfils nutritional needs of older Canadians is currently unknown; no evidence from contemporary eating patterns exists. The general aim of this study was therefore to assess the relationship between adherence to CFG-2019 recommendations, using the Healthy Eating Food Index-2019 (HEFI-2019), and intakes of key nutrients from food sources only, of adults aged ≥65 years from the CCHS 2015 ‒ Nutrition. The HEFI-2019 score is a metric measuring the alignment of eating patterns with CFG-2019 recommendations on food choices (17, 18). More precisely, we assessed 1) the continuous relationship between the HEFI-2019 score and nutrient intakes, and 2) the estimated change in the prevalence of nutrient inadequacy according to a hypothetical increase in HEFI-2019 score. We hypothesized that higher adherence to CFG-2019 recommendations would show an inverse relationship with intakes of nutrients more commonly found or eaten in animal-based foods, such as protein, calcium, vitamin D, iron, zinc, vitamin B6 and B12, and a positive relationship with intakes of nutrients more commonly found in plant-based foods such as fibre, folate, magnesium, potassium, vitamin A.

## Methods

### Study design and participants

This study is based on a sample of older adults aged 65 years or older from the CCHS 2015 – Nutrition (19). The CCHS 2015 -Nutrition is a nationally representative survey of individuals aged 1 year and older living in private dwellings in the 10 Canadian provinces. Full-time members of the Canadian Forces and individuals living in the Territories, on reserves, in remote areas, and in institutions were not included. Data collection occurred between January 1st to December 31, 2015. Respondents aged less than 65 years (n=16,394) and those reporting zero energy on their first 24-hour dietary recall intake were excluded (n=4), yielding a final sample of 4,089 respondents. The public use microdata file (PUMF) of CCHS 2015 – Nutrition was obtained from Statistics Canada.

### Data collection and dietary assessment

Interviewer-administered 24-hour dietary recalls were used to assess dietary intakes. The interviews were mostly (98%) conducted in person for the first 24-hour recall and by telephone for the second 24-hour recall. All 24-hour recall interviews were structured according to the Automated Multiple Pass Method (19). Portion size estimation of foods and beverages in plates, bowls, glasses and mugs was facilitated using a food booklet designed for the survey (19). All respondents completed one 24-hour recall and a subsample of 1,706 respondents (42%) completed a second 24-hour recall. Nutrient intakes were calculated based on the Canadian Nutrient File 2015 (20), except for intakes of free sugars which were recently published by Health Canada (21). Total food intakes expressed in reference amounts (RA) (22) were calculated for each respondent, 24-hour dietary recall and HEFI-2019 food and beverage categories. The interviewers measured body weight of respondents with a standard scale (LifeSource Scales Model US-321).

#### The Healthy Eating Food Index (HEFI) 2019

The HEFI-2019 is a continuous score which measures the degree of adherence between dietary intakes and Canada’s Food Guide 2019 recommendations on healthy food choices. Complete details about the development and the evaluation of the HEFI-2019 are available elsewhere (17, 18). Briefly, the HEFI-2019 has 10 components including 5 based on intake of foods (*Vegetables and fruits, Whole-grain foods, Grain foods ratio, Protein foods, Plant-based protein foods*), 1 on beverages (*Beverages*), and 4 on nutrients (*Fatty acids ratio, Saturated fats, Free sugars, and* Sodium). The *Saturated fats, Free sugars and Sodium* components are so-called “moderation” components, for which higher scores are attributed to intakes below thresholds consistent with CFG-2019 recommendations. The remaining components are “adequacy” components for which higher scores are attributed to greater intakes. Details about each component and scoring standards are presented in **Supplemental Table 1**. The total HEFI-2019 score sums up to 80 points with higher scores indicating greater adherence to recommendations. The HEFI-2019 was evaluated using dietary intake data from the CCHS 2015 – Nutrition (18). Notably, the variability of the HEFI-2019 score was sufficient, the HEFI-2019 score was correlated with the Healthy Eating Index (HEI)-2015 (r=0.79) and differences of HEFI-2019 score were found among subgroups with known diet quality differences based on sex, age, and smoking status (18). Finally, a prospective analysis of UK Biobank data suggested that a hypothetical increase in HEFI-2019 score reduces the 11-year risk of cardiovascular disease in middle-aged and older adults (6).

#### Nutrient intakes

Key nutrients for older adults were selected *a priori* according to data availability in the CCHS 2015 – Nutrition as well as previous literature (13, 23). The nutrients were protein, calcium, vitamin D, iron, zinc, vitamin B6, vitamin B12, dietary folate equivalent (folate hereafter), magnesium, potassium, fibre, and retinol activity equivalent (vitamin A hereafter). Nutrient intakes from supplements were not included since Canada’s Food Guide recommendations on healthy food choices focus on foods and beverages. The proportion of respondents with inadequate intakes was estimated using the cut-point method and the Estimated Average Requirements (EAR) of the Dietary Reference Intake (24). Because there is no EAR for potassium and fibre, the prevalence of intake inadequacy cannot be assessed. Instead, the proportion of respondents with intakes above the adequate intake (AI) value was considered.

Recognizing the consensus that recommended protein intakes for older adults should be higher than current recommendations (25–27), we also estimated the proportion of respondents with inadequate protein intakes at cut-offs higher than the current EAR. The hypothetical EAR cut-offs of 0.8 and 1.0 grams of protein per kg of bodyweight were selected to reflect hypothetical higher recommended daily allowance (RDA) of 1.0 and 1.2 g/kg, respectively. The latter analysis was not pre-specified and is considered exploratory.

### Statistical analyses

Sampling weights provided by Statistics Canada were used in all analyses to reflect the Canadian population of older adults in 2015 as well as bootstrap replicate weights for variance estimation. The sampling weights accounting for missing data on body weight were also used where appropriate. Analyses were performed in SAS Studio v3.8 (SAS Institute) and R v4.2.2 (R Foundation for Statistical Computing).

To assess the relationship between the HEFI-2019 score and nutrient intakes, analyses involved four steps, detailed below: 1) measurement error correction of dietary intakes to estimate usual intakes using the National Cancer Institute (NCI) Markov Chain Monte Carlo (MCMC) multivariate method; 2) linear regression of continuous nutrient intakes on total HEFI-2019 scores; 3) logistic regression of the proportion of individuals below EAR on total HEFI-2019 scores; and 4) variance estimation using bootstrap replicate weights provided by Statistics Canada.

First, the 24-hour dietary recalls are mainly affected by within-individual random errors, which require correction to estimate distribution of intakes (28, 29). To account for correlated random errors of all dietary constituents of the HEFI-2019 (e.g., vegetables and fruits, free sugars) and those of nutrient intakes (e.g., protein), the NCI MCMC multivariate method was used (MULTIVAR_MCMC_MACRO_V2.1 and MULTIVAR_DISTRIB_MACRO_V2.1, 2017) (30, 31). Briefly, the multivariate method uses Monte Carlo simulation to estimate distribution of usual intakes (i.e., long-term average) of multiple dietary constituents correlated with each other. The multivariate method also accounts for systematic differences due to “nuisance” factors (day of the week, sequence of recall) and considers foods that are episodically consumed (e.g., plant-based protein foods) (30).

Nutrients with common food sources were modelled together to have parsimonious measurement error correction models, yielding 5 different models each combining 1 to 4 nutrients with the 15 dietary constituents of the HEFI-2019 (see below). Model 1 included protein only; model 2 included calcium and vitamin D; model 3 included iron, zinc, vitamin B6 and vitamin B12; model 4 included folate, magnesium, potassium and fibre; model 5 included vitamin A only. The 15 dietary constituents of the HEFI-2019 were vegetables and fruits, whole-grain foods, animal-based protein foods, plant-based protein foods, unsweetened milk, water and other healthy beverages (e.g., unsweetened coffee or tea), refined grain foods, “other” foods not recommended, “other” beverages not recommended (i.e., sugary drinks, artificially sweetened beverages, vegetable and fruit juices, sweetened milk and plant-based beverages, alcohol), free sugars, monounsaturated fats, polyunsaturated fats, saturated fats, sodium and total energy intake. A total of 5 food and beverage categories (i.e., whole-grain foods, refined grain foods, plant-based protein foods, “other” beverages not recommended and unsweetened milk) were considered episodically consumed since at least 10% of respondents reported zero consumption on the first 24-h recall (31). All the remaining foods and nutrients were considered as consumed daily.

The measurement error correction models were stratified by sex to reflect sex-specific random variations in dietary intakes and to derive sex-specific associations (31, 32). The models also included the covariates age (indicator for 71 years or older), sequence of recall (indicator for second recall) and weekend (indicator for a 24-hour recall of intakes on Friday, Saturday or Sunday). In addition, the model for protein intake also included body weight (kg) as a covariate to derive protein intake per kg and assess the proportion of individuals below the EAR. A total of 500 pseudo-individuals were generated in the Monte Carlo simulation step of the NCI multivariate method. Simulations from each stratum (i.e., males and females) were pooled together. The HEFI-2019 scoring algorithm was then applied using the modelled dietary constituents among the 500 pseudo-individuals to derive HEFI-2019 scores. Descriptive statistics were calculated based on usual intakes among the 500 pseudo-individuals including the prevalence of intake inadequacy, mean (SD) dietary intakes and HEFI-2019 scores as well as correlations between dietary constituents of the HEFI-2019 and nutrient intakes.

Second, based on the Monte Carlo simulation data, simple linear regression models were used to assess the relationship between the continuous HEFI-2019 score, as the independent variable, and continuous nutrient intakes, as the dependent variable. A restricted cubic spline transformation with 5 knots (percentiles 5, 27, 50, 73 and 95) was applied *a priori* to the HEFI-2019 score to assess potential non-linearity (33). To estimate change according to feasible increases in adherence, expected nutrient intake differences were calculated according to an increase in HEFI-2019 score from the median score to the 90 ^th^ percentile of the usual intake distribution. In other words, we estimated nutrient intake difference according to a hypothetical change where respondents would have had high HEFI-2019 scores compared with the HEFI-2019 score respondents had on average, taken as the median of this sample.

Third, also based on the Monte Carlo simulation data, logistic regression models were used to assess the odds of having nutrient intake below the age- and sex-specific EAR and the continuous HEFI-2019 score. A restricted cubic spline transformation was also applied to the HEFI-2019 score. Predicted odds of nutrient intake inadequacy were generated for the 90^th^ percentile and the median. Both predicted odds were then re-expressed as risk of inadequacy inline1; where *X* corresponds to predicted odds at a given HEFI-2019 score percentile). The expected change in the prevalence of inadequate nutrient intakes, i.e., risk difference, was calculated by the difference between the estimated risk of inadequacy at the 90 ^th^ percentile vs. the median HEFI-2019 score.

Fourth, steps 1 to 3 were repeated 500 times using bootstrap replicate weights to generate standard errors and 95%CI via normal approximation. The convergence of bootstrap standard errors and the normality of bootstrap estimates were confirmed graphically. Data from one bootstrap replicate was removed for potassium due to non-convergence.

## Results

### HEFI-2019 score and dietary constituents

The mean (SD) HEFI-2019 score among all adults 65 years and older was 46.0 (8.9) (/80 points). Among age and sex subgroups, the mean HEFI-2019 score was the highest in females, 65 to 70 years (48.2 points) and the lowest in males, 65 to 70 years (44.9 points; **Figure 1**).

**Figure 1:**
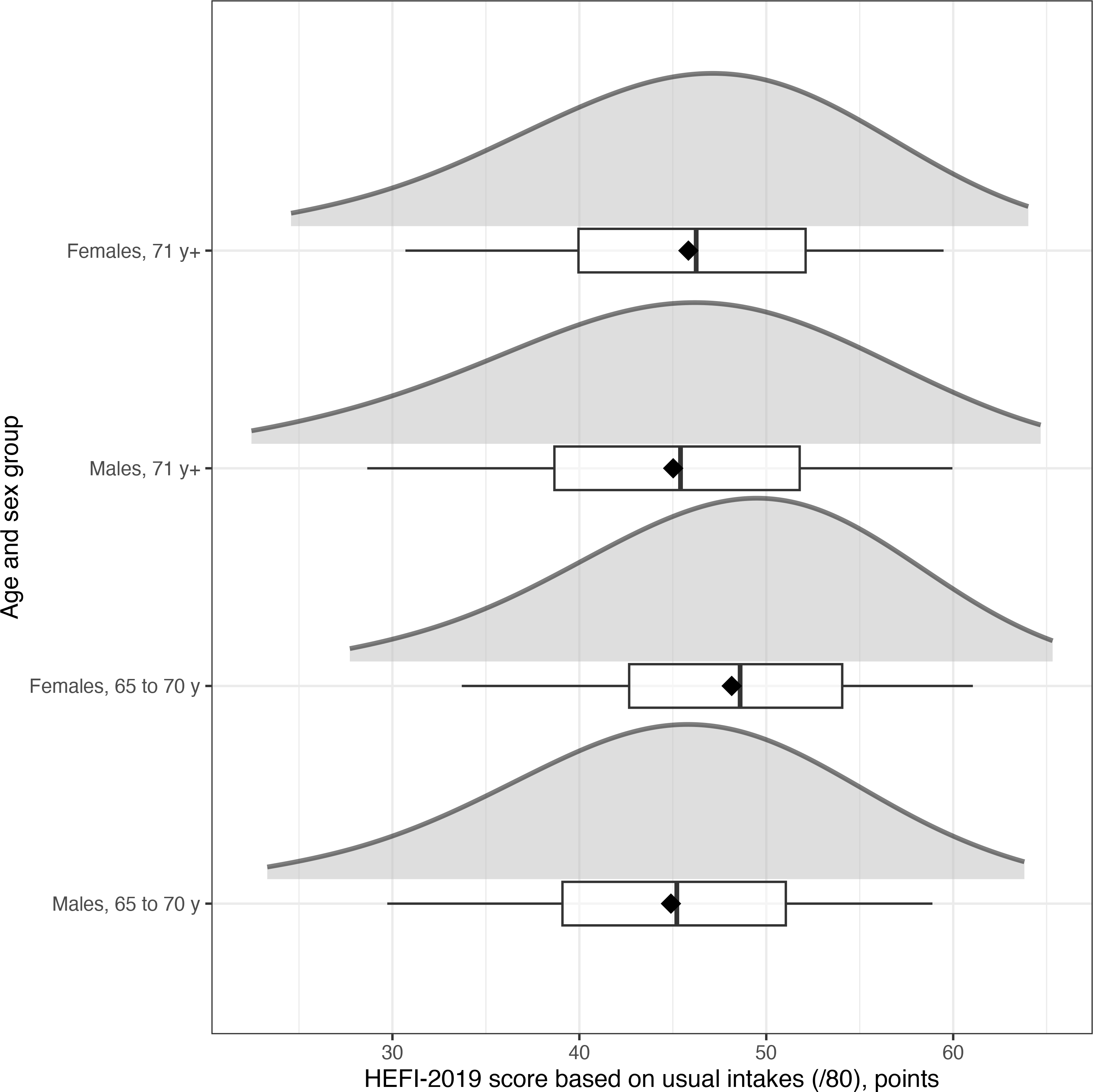
Distribution of total HEFI-2019 score, based on usual dietary intakes in 4,089 adults aged 65 years and more from the CCHS 2015 - Nutrition. Diamonds are means. Left and right whiskers indicate the 5th and 95th percentile, respectively. Dietary intakes were modelled using the National Cancer Institute multivariate method to estimate usual intakes (see Methods). CCHS, Canadian Community Health Survey; DRI, Dietary Reference Intake.

**Supplemental Table 2 and 3** present, respectively, usual mean intakes and percentile of the distribution of foods and beverages, as well as nutrients, contributing to the HEFI-2019 score. Older adults consumed approximately 84% of their total protein foods from animal-based source and 65% of their total grain foods intake as non-whole grain foods.

**Supplemental Figure 1** presents Pearson correlations between food and beverage categories contributing to the HEFI-2019 score and key nutrients. Most food and beverage categories recommended in CFG 2019 were correlated with greater nutrient intakes. Notably, the highest correlations were observed between milk and calcium (r=0.56), milk and vitamin D (r=0.54), vegetables and fruits and fibre (r=0.54) as well as whole-grain foods and fibre (r=0.54). Among foods and beverages not recommended in CFG, non-whole grain foods was the only group showing positive correlations with nutrient intakes including with folate (r=0.16) and with vitamin D (r=0.12).

### Prevalence of inadequate nutrient intake

The prevalence of inadequate nutrient intakes ranged from 1% (iron) to 96% (vitamin D; **Figure 2**. The prevalence of inadequate intakes was high for vitamin D, calcium, magnesium (96%, 83%, 64%, respectively), and moderately high for vitamin B6, vitamin A, folate and zinc (38%, 36%, 30%, 28%, respectively); Figure 1. Results were similar for most nutrients when stratified by DRI age and sex group (**Supplemental Table 4**).

**Figure 2:**
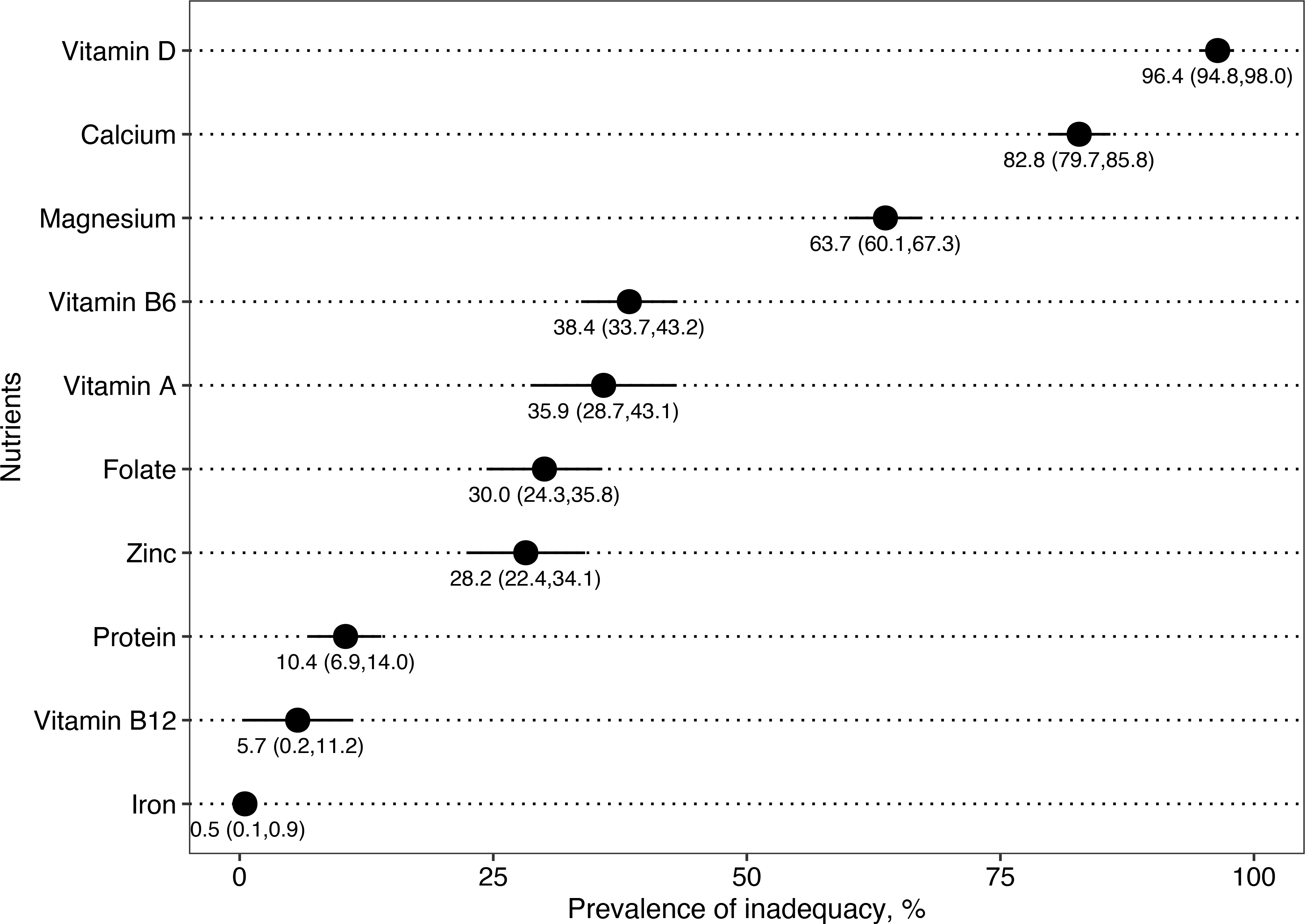
Prevalence of inadequate nutrient intakes in 4,089 adults aged 65 years and older from the CCHS 2015 - Nutrition. Only nutrient intakes from foods were considered (i.e., excluding intakes from dietary supplement). Inadequate intakes are intakes below the age- and sex-specific Estimated Average Requirements (EAR). Potassium and fibre are not shown, because only Adequate Intakes (AI) values are available for these nutrients. All dietary intakes were modelled using the National Cancer Institute multivariate method to estimate usual intakes (see Methods). 95%CI were estimated using 500 bootstrap weight replicates. CCHS, Canadian Community Health Survey.

### Relationship between HEFI-2019 score and nutrient intakes

Relationships between the HEFI-2019 score and nutrient intakes are illustrated in **Figure 3**. Respondents with higher HEFI-2019 score had higher intakes of fibre, magnesium, vitamin B6, potassium, and protein, but lower intakes of vitamin D, vitamin B12, dietary folate equivalent, and iron. The associations between the HEFI-2019 score and intakes of vitamin A, calcium, and zinc were weak or null. **Table 1** presents the expected differences in nutrient intakes associated with an increase in HEFI-2019 score to the 90^th^ percentile of the score distribution compared with median HEFI-2019 score. For example, increasing HEFI-2019 from the median to the 90^th^ percentile of the score distribution was associated with a 3.4 g/day higher fiber intake (95%CI: 2.0, 4.8; Table 1). In contrast, the same HEFI-2019 score increase was associated with a 0.5 μg/day lower vitamin D intake (95%CI: -1.0, 0.0; Table 1).

**Figure 3:**
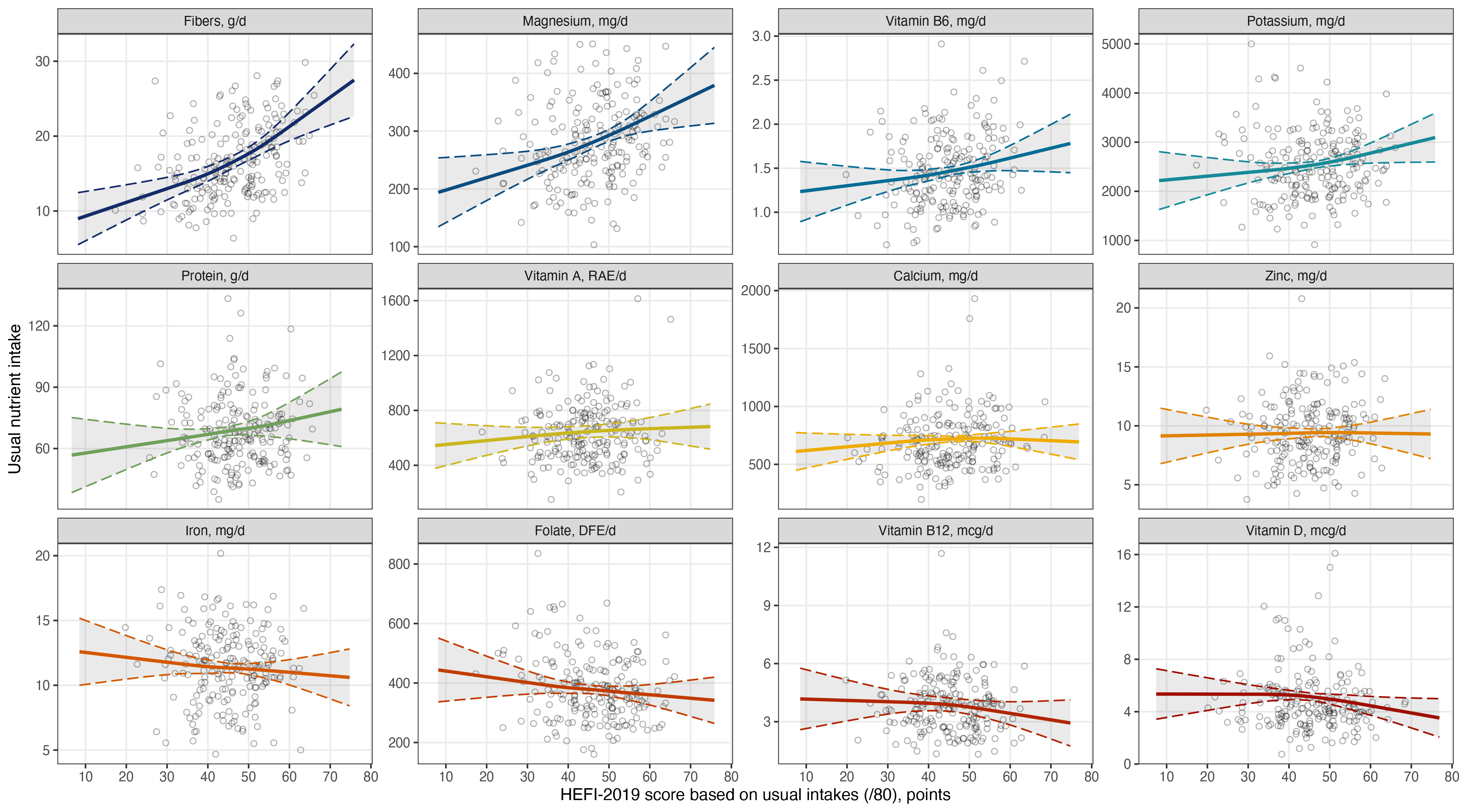
Linear regression of nutrient intake on the total HEFI-2019 score in 4,089 adults aged 65 years or more from the CCHS 2015 - Nutrition. A positive relationship indicates that greater HEFI-2019 scores are associated with greater nutrient intake, and inversely. For visualization purpose, data points are from a random sample of 200 respondents selected proportionally to sampling weights. All dietary intakes were modelled using the National Cancer Institute multivariate method to estimate usual intakes (see Methods). 95%CI were estimated using 500 bootstrap weight replicates. CCHS, Canadian Community Health Survey; HEFI-2019, Healthy Eating Food Index-2019.

**Table 1.** Usual nutrient intake difference between HEFI-2019 scores at the 90^th^ percentile and the median HEFI-2019 score distribution in 4,089 adults aged 65 years or more from the CCHS 2015 - Nutrition.

| Nutrients<sup>1</sup> | Dietary Reference Intake group |  |  |  |  |
| --- | --- | --- | --- | --- | --- |
|  | Adults,<br>65 y+<br>(n=4,089) | Males,<br>65 to 70 y<br>(n=567) | Females,<br>65 to 70 y<br>(n=720) | Males,<br>71 y+<br>(n=1,246) | Females,<br>71 y+<br>(n=1,556) |
| Protein, g/d | 3 (-2, 8) | 2 (-4, 9) | 5 (-1, 11) | 2 (-4, 8) | 4 (-2, 10) |
| Calcium, mg/d | 4 (-50, 57) | -16 (-96, 65) | 21 (-47, 89) | -14 (-88, 60) | 25 (-38, 89) |
| Vitamin D, mcg/d | -0.5 (-1.0, 0.0) | -0.8 (-1.6, -0.1) | -0.1 (-0.7, 0.5) | -0.7 (-1.5, 0.0) | -0.1 (-0.7, 0.6) |
| Iron, mg/d | -0.2 (-1.0, 0.5) | -0.6 (-1.8, 0.6) | 0.2 (-0.7, 1.1) | -0.6 (-1.8, 0.6) | 0.3 (-0.5, 1.1) |
| Vitamin B12, mcg/d | -0.3 (-0.7, 0.1) | -0.4 (-0.9, 0.0) | -0.1 (-0.7, 0.4) | -0.4 (-0.8, 0.1) | -0.1 (-0.6, 0.4) |
| Vitamin B6, mg/d | 0.1 (0.0, 0.2) | 0.1 (0.0, 0.2) | 0.1 (0.0, 0.3) | 0.1 (0.0, 0.2) | 0.1 (0.0, 0.2) |
| Zinc, mg/d | 0.0 (-0.7, 0.7) | -0.4 (-1.5, 0.7) | 0.4 (-0.3, 1.2) | -0.3 (-1.3, 0.6) | 0.5 (-0.2, 1.2) |
| Folate, mcg/d | -13 (-39, 14) | -15 (-56, 26) | -6 (-36, 25) | -16 (-55, 22) | -2 (-31, 27) |
| Fibre, g/d | 3 (2, 5) | 4 (2, 7) | 3 (1, 5) | 4 (2, 6) | 3 (1, 5) |
| Magnesium, mg/d | 33 (14, 52) | 44 (14, 73) | 30 (6, 55) | 39 (13, 65) | 31 (9, 53) |
| Potassium, mg/d | 173 (17, 328) | 230 (6, 455) | 170 (-27, 368) | 214 (3, 424) | 178 (-7, 363) |
| Vitamin A, RAE/d | 16 (-37, 69) | 9 (-54, 71) | 26 (-54, 106) | 10 (-48, 68) | 26 (-42, 95) |

<sup>1</sup>Values are expected nutrient intake differences (95%CI) for a HEFI-2019 score at the 90<sup>th</sup> compared with the 50<sup>th</sup> percentile of the score distribution, and were estimated using linear regression models. Estimates reflect the expected nutrient intake difference when HEFI-2019 scores are increased to correspond to the 90<sup>th</sup> percentile of the score distribution, compared with the median HEFI-2019 score. All dietary intakes were modelled using the National Cancer Institute multivariate method to estimate usual intakes (see Methods). 95%CI were estimated using 500 bootstrap weight replicates. Sample size indicate the number of respondents without applying sampling weights, but other estimates were weighted to reflect the Canadian population. CCHS, Canadian Community Health Survey; d, day; HEFI-2019, Healthy Eating Food Index-2019.

## Relationship between the HEFI-2019 score and nutrient intake inadequacy

### Inadequate nutrient intakes

**Figure 4** presents the prevalence and the difference in the prevalence of inadequate nutrient intake according to the HEFI-2019 score. An increase of HEFI-2019 score to the 90^th^ percentile of the score distribution compared with median HEFI-2019 score was associated with reduction of 20, 13, and 5 percentage points in the prevalence of inadequate nutrient intakes for magnesium, vitamin B6, and protein, respectively. Inversely, the same HEFI-2019 score increase was associated with a minor increase in the prevalence of inadequate vitamin D intakes (prevalence difference, +2.6%; 95%CI: 1.1, 4.0%). The prevalence of inadequate iron intake was 0% and thus unaffected by hypothetical HEFI-2019 score change. For vitamin A, zinc, vitamin B12, calcium and folate, the 95%CI were wide and compatible with both increase and decrease in the prevalence of inadequate intakes (Figure 3).

**Figure 4:**
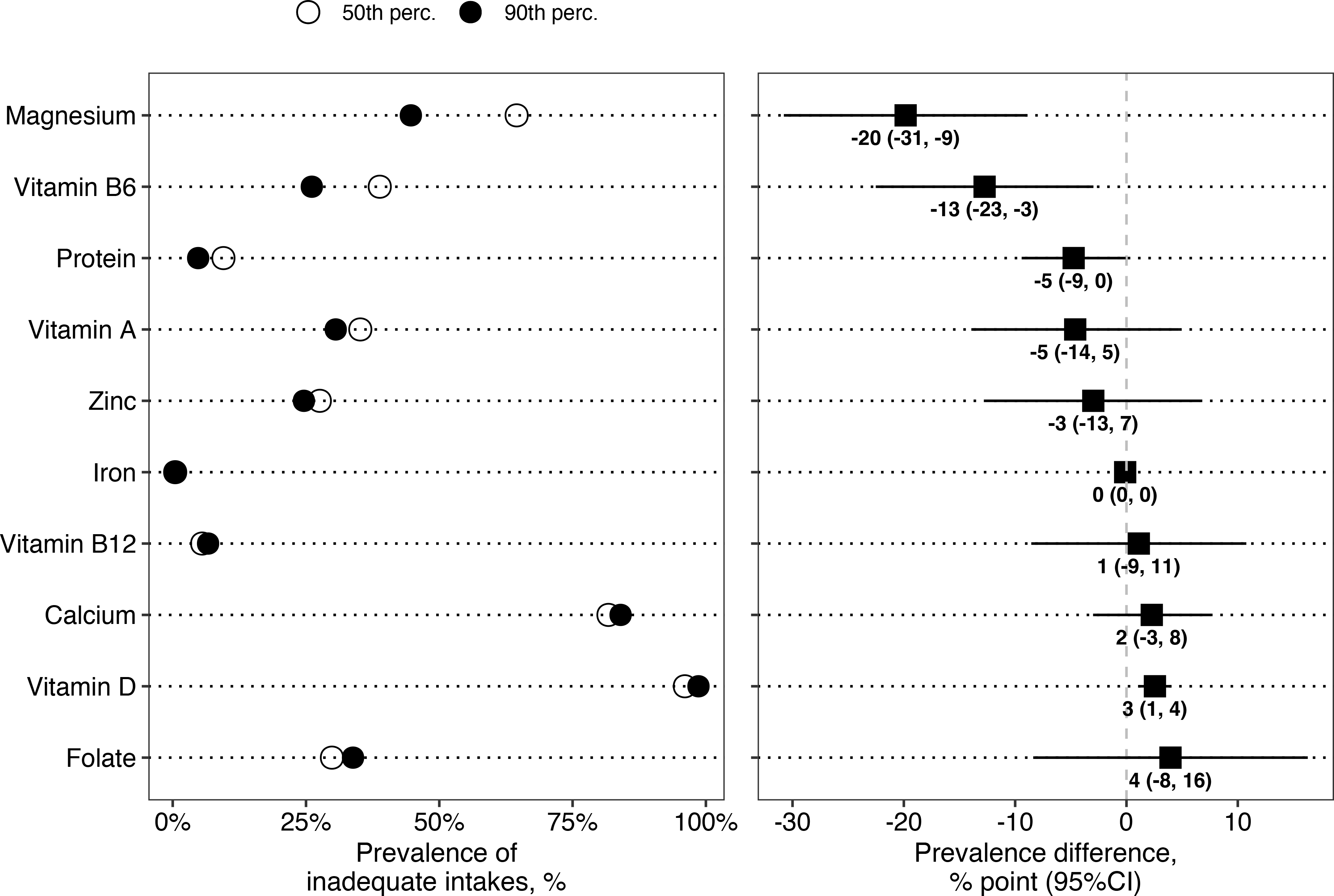
Prevalence of inadequate intakes and difference for HEFI-2019 scores at the 90th compared with the 50th percentile of the score distribution in 4,089 adults aged 65 years or more from the CCHS 2015 - Nutrition. Inadequate intakes are intakes below the age- and sex-specific Estimated Average Requirements (EAR). Potassium and fibre are not shown, because only Adequate Intakes (AI) values are available for these nutrients. All dietary intakes were modelled using the National Cancer Institute multivariate method to estimate usual intakes (see Methods). 95%CI were estimated using 500 bootstrap weight replicates. CCHS, Canadian Community Health Survey; HEFI-2019, Healthy Eating Food Index-2019.

#### Hypothetical higher Estimated Average Requirements for protein

Compared with the median HEFI-2019 score, scores at the 90^th^ percentile decreased the proportion of respondents with protein intakes below hypothetical EAR of 0.8 g/kg/day and 1.0 g/kg/day (**Supplemental Figure 2**). For 0.8 g/kg/day, the prevalence of inadequacy decreased from 29.3% to 18.5% (prevalence difference: -10.8; 95%CI: -20.0, -1.6). For 1.0 g/kg/day, the prevalence decreased from 64.7% to 50.8% (prevalence difference: -13.9; 95%CI: -25.0, -2.8).

### Adequate intakes for fibre and potassium

Compared with the median HEFI-2019 score, score at the 90^th^ percentile increased the proportion of respondents with intakes above the adequate intake for both fibre and potassium. For fibre, the prevalence increased from 6.5% to 20.5% (prevalence difference: +14.1; 95%CI: 5.1, 23.0). For potassium, the prevalence increased from 25.1% to 36.8% (prevalence difference: +11.7; 95%CI: 2.2, 21.1).

## Discussion

The objective of this study was to describe the relationship between adherence to CFG-2019 recommendations on healthy food choices, measured using the HEFI-2019, and nutrient intakes from food sources in adults 65 years and older from Canada in 2015. We found that higher adherence was associated with higher intakes of fibre, magnesium, vitamin B6, potassium and protein. Had respondents had a higher adherence, we estimated that the prevalence of nutrient inadequacy from foods and beverages for magnesium, vitamin B6 and protein would have been 20%, 13% and 5% lower, respectively. However, a higher adherence was not associated with higher intakes of calcium, zinc, iron, folate, vitamin B12 and vitamin D. In turn, we estimated that the high prevalence of calcium inadequacy would not have changed and that of food-based vitamin D would have been 3% higher in 2015. Overall, these results indicate that CFG-2019 recommendations are insufficient to mitigate nutrient intake inadequacy for certain key nutrients based on the eating patterns of adults aged 65 years or more from Canada in 2015. These findings partially confirm our hypothesis that nutrients typically found in animal-based foods (e.g., iron, vitamin B12, calcium, vitamin D) were inversely associated with adherence to CFG.

Few studies have examined nutrient intake inadequacy according to adherence to the CFG-2019 recommendations. Barr (9) assessed the probability of intake inadequacy upon adherence to an eating pattern consistent with foods depicted in the CFG-2019 plate snapshot. The probability of inadequacy was near 100% for calcium and vitamin D for males and females aged 71 years or more. This result is consistent with findings in the present study, where a higher adherence to recommendations on healthy food choices was insufficient to mitigate the prevalence of inadequacy for calcium and vitamin D. The lack of specific recommendations regarding dairy foods and alternatives in CFG-2019, and consequently the absence of points in the HEFI-2019, may partly explain these findings. Among food and beverage categories contributing to the HEFI-2019, unsweetened milk had the highest correlation with intakes of calcium (r=0.56) and vitamin D (r=0.53; Supplemental Figure 1). Indeed, the main food source of calcium and vitamin D in Canadians’ diet was the “Milk & Alternatives” food group in 2015 (34–36). Of note, the prevalence of inadequacy for vitamin D was very high in this sample (>95%), supporting the notion that meeting vitamin D recommendations through foods alone is an important challenge (36). Similarly, data from the CCHS indicate that calcium intake inadequacy in supplement non-users increased from 58% to 68% between 2004 and 2015, respectively (35). All in all, higher adherence to CFG-2019 recommendations on food choices was insufficient to mitigate the high prevalence of inadequate calcium and vitamin D intakes. Thus, meeting calcium and vitamin D needs would require additional strategies such as specific recommendations in CFG in addition to supplementation or food fortification.

A review by Fernandez et al. highlighted potential gaps in the application of CFG-2019 recommendations, notably whether the CFG-2019 permitted the adequate consumption of nutrient-rich protein foods or not (10). Our results revealed that a higher adherence to CFG-2019 recommendations slightly reduced the low proportion of older adults with intakes below the current EAR (0.66 g/kg) and reduced by 11% and 14% the proportion of those below the hypothetical EAR of 0.8 g/kg and 1.0 g/kg. Notwithstanding, the prevalence of inadequacy for the hypothetical EAR would remain considerable, with 25% and 50% inadequate protein intakes, respectively, thus representing an additional risk factor for sarcopenia (26). Furthermore, these findings only reflect the total daily protein intake and the impact on overall protein food quality or distribution during the day is unknown. Future studies should investigate the relationship between higher adherence to CFG-2019 and overall protein food quality and should examine how to increase consumption of high-quality plant-based protein foods. In that regard, a qualitative study among older adults highlighted that health benefits and food preparation skills would facilitate consumption of plant-based protein foods (37). Increasing consumption of unsweetened soy beverage (currently zero in the present study) or unsweetened milk (mean intake of 125 ml/d; Supplemental Table 2) could also contribute to higher intake of high-quality protein. Finally, we stress that the CCHS 2015 – Nutrition excluded individuals living in institutions who likely have additional needs as well as barriers to consuming higher protein diets (14).

The inverse associations between the HEFI-2019 and intakes of iron, folate, and vitamin B12 also raise concerns. On one hand, the prevalence of inadequacy for these nutrients did not increase at the population level, partly because the EAR can be met at relatively low intakes for these nutrients. On the other hand, at the individual level, meeting nutrient requirements while following CFG-2019 recommendations may be more difficult in individuals at risk of inadequate intakes, e.g., older adults with loss of appetite (13, 14). Further, vitamin B12 may be particularly important for older adults and recent evidence support that food groups contribute differently at reducing risk of deficiency (38). Meeting folate requirements while following CFG may also be challenging for some individuals since refined grains, not recommended in CFG, showed the highest correlations with folate intake in the present study (Supplemental Figure 1). Indeed, non-whole grain foods are subject to mandatory fortification in Canada which is not the case of whole grain foods (39). In sum, CFG-2019 recommendations should be carefully implemented to ensure meeting nutrient intakes both at the population and individual level. Health Canada published additional guidance in 2022 on how to apply dietary guidelines to support health professionals and policy (40). The guidance describes the type and frequency of foods to support nutritional needs. The extent to which the additional guidance helps older adults shift their eating patterns to be both consistent with CFG-2019 and provide optimal nutrient intakes remains to be determined. Findings of the present study highlight that eating patterns of older adults in 2015 failed to provide optimal intakes of calcium, folate and vitamin D despite adhering to CFG-2019 recommendations thereby supporting the need for additional guidance, perhaps with quantitative recommendations.

The use of national survey data to estimate the impact of higher adherence on the prevalence of nutrient inadequacy in Canada is a strength of this study. But it must be recognized that relationships between adherence to CFG-2019 recommendations and nutrient intakes reflect the underlying eating patterns of adults 65 years or older from Canada in 2015. A variety of underlying food and beverage consumption patterns may be highly consistent with CFG-2019 recommendations yet, have diverging associations with intakes of certain nutrients. As well, eating patterns of older adults, though generally stable, may evolve over time which would result in different associations between adherence to CFG-2019 and nutrients intakes. Another key strength of this work is the use of the NCI MCMC multivariate method to account for random errors in dietary intakes measured with 24-hour recall (30, 31). Using the multivariate method permitted the joint assessment of the relationship between usual nutrient intakes and adherence to CFG-2019 recommendations on healthy food choices, as measured with the HEFI-2019, and the modelling of higher adherence to CFG-2019 recommendations compared with average adherence. Limitations must be addressed. First, self-reported dietary intakes with 24-hour dietary recall are affected by systematic errors or bias (41, 42). The working assumption that intakes are unbiased cannot be verified. Of note, we did not adjust for “misreporting status” to account for energy under-reporting in the present study. The proportion of under-reporters among older adults is not higher than in other age and sex group samples of the CCHS 2015 – Nutrition (43) and evidence indicates that diet quality based on 24-hour dietary recall may not be prone to large bias (44). Second, the food composition database, the Canadian Nutrient File 2015, may not adequately reflect the nutrient profile of foods actually consumed (45). Accordingly, estimates of prevalence of nutrient inadequacy based on self-reported dietary intakes data are not as accurate as estimates based on biomarker data. Third, our results cannot be used to extrapolate nutrient intake inadequacy at the national level. Although calcium and vitamin D supplements could contribute to reducing inadequacies (35, 36), nutrient intakes from dietary supplements were not included since the objective was to assess the relationship between nutrients and adherence to CFG-2019 recommendations on healthy food choices (4, 5, 17).

In conclusion, higher adherence to CFG-2019 recommendations on healthy food choices was associated with greater intakes of protein, fiber, magnesium and many nutrients considered, based on eating patterns of adults aged 65 years or more from the CCHS 2015 – Nutrition. However, higher adherence was associated with lower intakes of iron, folate, vitamin B12 and vitamin D, and not associated with calcium, zinc or vitamin A intakes, thus insufficient to mitigate dietary inadequacy in these key nutrients. Knowledge of shortfalls of CFG-2019 recommendations regarding nutrient intake inadequacy are relevant to help government, policymakers and registered dietitians in providing more comprehensive and appropriate recommendations to older adults. Our findings support the need for additional guidance, as that recently published by Health Canada to help ensure that eating patterns are both consistent with CFG-2019 and adequate nutrient intakes (40). Future studies should investigate how eating patterns changed after the publication of CFG-2019 and the additional guidelines and how these changes may affect the relationship between adherence to recommendations and nutrient intakes.

## Data Availability

The Canadian Community Health Survey 2015 Nutrition Public Use Microdata File are available upon request at https://www150.statcan.gc.ca/n1/en/catalogue/82M0024X2018001. The analytic code and data produced will be made freely available at https://github.com/didierbrassard.

## Abbreviations list

CCHS: Canadian Community Health Survey
CFG: Canada’s Food Guide
DRI: Dietary Reference Intake
HEFI-2019: Healthy Eating Food Index-2019
NCI: National Cancer Institute.

## Data availability statement

The Canadian Community Health Survey 2015 – Nutrition Public Use Microdata File are available upon request at https://www150.statcan.gc.ca/n1/en/catalogue/82M0024X2018001. The analytic code will be made freely available at https://github.com/didierbrassard.

## Authors’ contributions to the manuscript

DB and SC designed the research; DB performed statistical analysis; DB wrote the first draft of the manuscript; DB and SC gave final approval and critically reviewed the manuscript.

## References

1. Health Canada. Eating Well with Canada’s Food Guide. Ottawa: Health Canada, 2007:1–6.

2. Katamay SW, Esslinger KA, Vigneault M, Johnston JL, Junkins BA, Robbins LG, et al. Eating well with Canada’s Food Guide (2007): development of the food intake pattern. Nutr Rev 2007;65:155–66.

3. Health Canada. Food, Nutrients and Health: Interim Evidence Update 2018 for Health Professionals and Policy Makers. Ottawa: Health Canada, 2019.

4. Health Canada. Canada’s food guide. Ottawa: Health Canada, 2019.

5. Health Canada. Canada’s Dietary Guidelines - for Health Professionals and Policy Makers. 2019.

6. Brassard D, Manikpurage HD, Thériault S, Arsenault BJ, Lamarche B. Greater adherence to the 2019 Canada’s Food Guide recommendations on healthy food choices reduces the risk of cardiovascular disease in adults: a prospective analysis of UK Biobank data. Am J Clin Nutr 2022;116:1748–58.

7. Asher KE, Doucet S, Luke A. Registered dietitians’ perceptions and use of the plant-based recommendations in the 2019 Canada’s Food Guide. J Hum Nutr Diet 2021;34:715–23.

8. Barco Leme AC, Laila A, Hou S, Fisberg RM, Ma DWL, Fisberg M, et al. Perceptions of the 2019 Canada’s Food Guide: a qualitative study with parents from Southwestern Ontario. Appl Physiol Nutr Metab 2021.

9. Barr SI. Is the 2019 Canada’s Food Guide Snapshot nutritionally adequate? Appl Physiol Nutr Metab 2019;44:1387–90.

10. Fernandez MA, Bertolo RF, Duncan AM, Phillips SM, Elango R, Ma DWL, et al. Translating “protein foods” from the new Canada’s Food Guide to consumers: knowledge gaps and recommendations. Appl Physiol Nutr Metab 2020;45:1311–23.

11. Auclair O, Burgos SA. Protein consumption in Canadian habitual diets: usual intake, inadequacy, and the contribution of animal- and plant-based foods to nutrient intakes. Appl Physiol Nutr Metab 2021;46:501–10.

12. Fabek H, Sanchez-Hernandez D, Ahmed M, Marinangeli CPF, House JD, Anderson GH. An examination of contributions of animal- and plant-based dietary patterns on the nutrient quality of diets of adult Canadians. Appl Physiol Nutr Metab 2021;46:877–86.

13. Shlisky J, Bloom DE, Beaudreault AR, Tucker KL, Keller HH, Freund-Levi Y, et al. Nutritional Considerations for Healthy Aging and Reduction in Age-Related Chronic Disease. Adv Nutr 2017;8:17–26.

14. Remond D, Shahar DR, Gille D, Pinto P, Kachal J, Peyron MA, et al. Understanding the gastrointestinal tract of the elderly to develop dietary solutions that prevent malnutrition. Oncotarget 2015;6:13858–98.

15. Millen BE, Abrams S, Adams-Campbell L, Anderson CA, Brenna JT, Campbell WW, et al. The 2015 dietary guidelines advisory committee scientific report: development and major conclusions. Adv Nutr 2016;7:438–44.

16. Keller HH, Lengyel C, Carrier N, Slaughter SE, Morrison J, Duncan AM, et al. Prevalence of inadequate micronutrient intakes of Canadian long-term care residents. Br J Nutr 2018;119:1047–56.

17. Brassard D, Elvidge Munene LA, St-Pierre S, Guenther PM, Kirkpatrick SI, Slater J, et al. Development of the Healthy Eating Food Index (HEFI)-2019 measuring adherence to Canada’s Food Guide 2019 recommendations on healthy food choices. Appl Physiol Nutr Metab 2022;47:595–610.

18. Brassard D, Elvidge Munene LA, St-Pierre S, Gonzalez A, Guenther PM, Jessri M, et al. Evaluation of the Healthy Eating Food Index (HEFI)-2019 measuring adherence to Canada’s Food Guide 2019 recommendations on healthy food choices. Appl Physiol Nutr Metab 2022;47:582–94.

19. Health Canada. Reference Guide to Understanding and Using the Data: 2015 Canadian Community Health Survey - Nutrition. 2017:1–91.

20. Health Canada. Internet: https://food-nutrition.canada.ca/cnf-fce/index-eng.jsp.

21. Rana H, Mallet M-C, Gonzalez A, Verreault M-F, St-Pierre S. Free Sugars Consumption in Canada. Nutrients 2021;13:1471.

22. Health Canada. Nutrition Labelling: Table of Reference Amounts for Food. Ottawa, 2016.

23. Dorrington N, Fallaize R, Hobbs DA, Weech M, Lovegrove JA. A Review of Nutritional Requirements of Adults Aged >/=65 Years in the UK. J Nutr 2020;150:2245–56.

24. Otten JJ, Hellwig JP, Meyers LD. Dietary reference intakes : the essential guide to nutrient requirements. Washington, D.C.: National Academies Press, 2006.

25. Phillips SM, Chevalier S, Leidy HJ. Protein “requirements” beyond the RDA: implications for optimizing health. Appl Physiol Nutr Metab 2016;41:565–72.

26. Bauer J, Biolo G, Cederholm T, Cesari M, Cruz-Jentoft AJ, Morley JE, et al. Evidence-based recommendations for optimal dietary protein intake in older people: a position paper from the PROT-AGE Study Group. J Am Med Dir Assoc 2013;14:542–59.

27. Traylor DA, Gorissen SHM, Phillips SM. Perspective: Protein Requirements and Optimal Intakes in Aging: Are We Ready to Recommend More Than the Recommended Daily Allowance? Adv Nutr 2018;9:171–82.

28. Dodd KW, Guenther PM, Freedman LS, Subar AF, Kipnis V, Midthune D, et al. Statistical methods for estimating usual intake of nutrients and foods: a review of the theory. J Am Diet Assoc 2006;106:1640–50.

29. Thompson FE, Kirkpatrick SI, Subar AF, Reedy J, Schap TE, Wilson MM, et al. The National Cancer Institute’s Dietary Assessment Primer: A Resource for Diet Research. J Acad Nutr Diet 2015;115:1986–95.

30. Zhang S, Midthune D, Guenther PM, Krebs-Smith SM, Kipnis V, Dodd KW, et al. A New Multivariate Measurement Error Model with Zero-Inflated Dietary Data, and Its Application to Dietary Assessment. Ann Appl Stat 2011;5:1456–87.

31. Kirkpatrick SI, Guenther PM, Subar AF, Krebs-Smith SM, Herrick KA, Freedman LS, et al. Using short-term dietary intake data to address research questions related to usual dietary intake among populations and subpopulations: Assumptions, statistical techniques, and considerations. J Acad Nutr Diet 2022;122:1246–62.

32. Herrick KA, Rossen LM, Parsons R, Dodd KW. Estimating Usual Dietary In take From National Health and Nutrition Examination Survey Data Using the National Cancer Institute Method. Vital Health Stat 2018;2:1–63.

33. Harrell FE. General Aspects of Fitting Regression Models. Edtion ed. Regression Modeling Strategies: With Applications to Linear Models, Logistic and Ordinal Regression, and Survival Analysis. Cham: Springer International Publishing, 2015:13-44.

34. Vatanparast H, Islam N, Shafiee M. Consumption of Milk and alternatives decreased among Canadians from 2004 to 2015: evidence from the Canadian community health surveys. BMC Nutr 2021;7:63.

35. Vatanparast H, Islam N, Patil RP, Shafiee M, Whiting SJ. Calcium Intake from Food and Supplemental Sources Decreased in the Canadian Population from 2004 to 2015. J Nutr 2020;150:833–41.

36. Vatanparast H, Patil RP, Islam N, Shafiee M, Whiting SJ. Vitamin D Intake from Supplemental Sources but Not from Food Sources Has Increased in the Canadian Population Over Time. J Nutr 2020;150:526–35.

37. Drolet-Labelle V, Laurin D, Bedard A, Drapeau V, Desroches S. Beliefs underlying older adults’ intention to consume plant-based protein foods: A qualitative study. Appetite 2022:106346.

38. Huang HH, Cohen AA, Gaudreau P, Auray-Blais C, Allard D, Boutin M, et al. Vitamin B-12 Intake from Dairy But Not Meat is Associated with Decreased Risk of Low Vitamin B-12 Status and Deficiency in Older Adults from Quebec, Canada. J Nutr 2022;152:2483–92.

39. Government of Canada. 2022. Internet: https://inspection.canada.ca/food-labels/labelling/industry/nutrient-content/reference-information/eng/1389908857542/1389908896254?chap=1 (accessed June 2023).

40. Health Canada. 2022. Internet: https://food-guide.canada.ca/en/guidelines (accessed June 2023).

41. Freedman LS, Commins JM, Moler JE, Willett W, Tinker LF, Subar AF, et al. Pooled results from 5 validation studies of dietary self-report instruments using recovery biomarkers for potassium and sodium intake. Am J Epidemiol 2015;181:473–87.

42. Freedman LS, Commins JM, Moler JE, Arab L, Baer DJ, Kipnis V, et al. Pooled results from 5 validation studies of dietary self-report instruments using recovery biomarkers for energy and protein intake. Am J Epidemiol 2014;180:172–88.

43. Garriguet D. Accounting for misreporting when comparing energy intake across time in Canada. Health Rep 2018;29:3–12.

44. Kirkpatrick SI, Dodd KW, Potischman N, Zimmerman TP, Douglass D, Guenther PM, et al. Healthy Eating Index-2015 Scores Among Adults Based on Observed vs Recalled Dietary Intake. J Acad Nutr Diet 2021;121:2233–41 e1.

45. Satija A, Yu E, Willett WC, Hu FB. Understanding nutritional epidemiology and its role in policy. Adv Nutr 2015;6:5–18.

