## Supplemental Material for "Relationship between adherence to the 2019 Canada’s Food Guide recommendations on healthy food choices and nutrient intakes in older adults"

### Supplementary Methods

#### The Healthy Eating Food Index (HEFI) 2019

**Supplemental Table 1** presents the HEFI-2019 (Brassard et al. 2022) components, points and thresholds for scores.

**Supplementary Table 1.** Healthy Eating Food Index (HEFI)-2019 components, points and standards for scoring

| **#** | **Component name** | **Measurement (ratio)** | **Maximum Points** | **Unit** | **Standard for minimum score** | **Standard for maximum score** |
| --- | --- | --- | --- | --- | --- | --- |
| 1 | *Vegetables and fruits^1^* | Total vegetables and fruits / Total foods*^2^* | 20 | RA/RA | No vegetables and no fruits | ≥ 0.50 |
| 2 | *Whole-grain foods* | Total whole-grain foods / Total foods*^2^* | 5 | RA/RA | No whole-grain foods | ≥ 0.25 |
| 3 | *Grain foods ratio^3^* | Total whole-grain foods / Total grain foods*^4^* | 5 | RA/RA | No whole-grain foods | = 1.0 |
| 4 | *Protein foods^5^* | Total protein foods / Total foods*^2^* | 5 | RA/RA | No protein foods | ≥ 0.25 |
| 5 | *Plant-based protein foods^6^* | Plant-based protein foods / Total protein foods | 5 | RA/RA | No plant-based protein foods | > 0.50 |
| 6 | *Beverages* | (Plain water including carbonated + unsweetened beverages) / Total beverages*^7^* | 10 | g/g | No water and no unsweetened beverages | = 1.0 |
| 7 | *Fatty acids ratio* | (Mono- + polyunsaturated fat) / Total saturated fat | 5 | g/g | ≤ 1.1*^8^* | ≥ 2.6*^9^* |
| 8 | *Saturated fats* | Total saturated fat / energy | 5 | %E (kcal/kcal) | ≥ 15%E*^10^* | < 10%E |
| 9 | *Free sugars* | Total free sugars / energy | 10 | %E (kcal/kcal) | ≥ 20%E*^10^* | < 10%E |
| 10 | *Sodium* | Total sodium / energy | 10 | mg / kcal | ≥ 2.0 | < 0.9*^11^* |

*^1^* All vegetables and fruits regardless of saturated fat, sodium or free sugar content; excludes fruit juice (i.e., considered as sugary drinks in CFG-2019).

*^2^* Total foods include all foods consumed as well as beverages considered in protein foods (i.e., unsweetened milk and unsweetened plant-based beverages that contain protein); excludes all other beverages as well as solid fats, oils and spreads and culinary ingredients (e.g., spices and baking soda).

*^3^* Foods where the first ingredient is either whole grains or whole wheat, regardless of saturated fat, sodium or free sugar content.

*^4^* Foods where the first ingredient is a grain (whole or not) regardless of saturated fat, sodium or free sugar content.

*^5^* All protein foods regardless of fat, sodium or sugars content; excludes processed meats (i.e., not considered protein foods in CFG-2019) and sweetened milks (i.e., considered as sugary drinks in CFG-2019).

*^6^* All plant-based protein foods, regardless of saturated fat, sodium or free sugar content.

*^7^* Unsweetened beverages include unsweetened coffee and tea, unsweetened milk and unsweetened plant-based beverages. Total beverages include water (plain or carbonated), coffee, tea, milk and plant-based beverages, fruit and vegetable juices, alcoholic drinks, artificially sweetened beverages and sugary drinks.

*^8^* Approximately the **15th percentile** of intake based on data (single 24-h dietary recall) in Canadians from the 2015 CCHS – Nutrition.

*^9^* Corresponds to the **1st percentile** of unsaturated to saturated fats ratios among simulated diets developed to be fully consistent with all recommendations in CFG-2019.

*^10^* Approximately the **85th percentile** of intake based on data (single 24-h dietary recall) in Canadians from the 2015 CCHS – Nutrition.

*^11^* Standard for maximum points based on the Chronic Disease Risk Reduction for 14+ years (i.e., 2300 mg) over the **90th percentile** of usual energy intakes in respondents 2 y and older from the 2015 CCHS – Nutrition (i.e., approximately 2600 kcal).

Table adapted from Brassard et al. Appl Physiol Nutr Metab. 2022. CCHS, Canadian Community Health Survey; CFG-2019, Canada's food guide 2019; HEFI-2019, Healthy Eating Food Index 2019; RA, Reference Amounts (amount of food usually eaten by an individual at one sitting); %E, percent of total energy.

### Supplementary Results

#### Usual food and nutrient intakes

**Supplemental 2 and 3** present estimated means and percentile of the distribution of foods, beverages and nutrients considered in the HEFI-2019 score.

**Supplemental Table 2.** Estimated means and percentiles of food and beverage categories considered in the HEFI-2019 score in adults aged 65 years or more from the CCHS 2015 - Nutrition*^1^*

|  |  | **Percentile** | | | | | | | | |
| --- | --- | --- | --- | --- | --- | --- | --- | --- | --- | --- |
| Categories | Mean (SD) | 1 | 5 | 10 | 25 | 50 | 75 | 90 | 95 | 99 |
| **Recommended in CFG** | | | | | | | | | | |
| Vegetables and fruits, RA/d | 3.5 (1.5) | 1.0 | 1.5 | 1.8 | 2.4 | 3.3 | 4.3 | 5.5 | 6.3 | 8.2 |
| Whole-grain foods, RA/d | 0.9 (0.6) | 0.0 | 0.1 | 0.1 | 0.4 | 0.8 | 1.3 | 1.7 | 2.0 | 2.5 |
| Protein foods, animal-based, RA/d | 2.1 (0.9) | 0.6 | 0.9 | 1.1 | 1.5 | 2.0 | 2.6 | 3.3 | 3.7 | 4.6 |
| Protein foods, plant-based, RA/d | 0.4 (0.3) | 0.0 | 0.1 | 0.1 | 0.2 | 0.3 | 0.6 | 0.9 | 1.1 | 1.6 |
| Milk, RA/d | 0.5 (0.5) | 0.0 | 0.0 | 0.1 | 0.2 | 0.4 | 0.8 | 1.2 | 1.6 | 2.4 |
| Water and healthy beverages, ml/d | 1,150 (563) | 238 | 402 | 513 | 741 | 1,064 | 1,461 | 1,897 | 2,196 | 2,855 |
| **Not recommended in CFG** | | | | | | | | | | |
| SSBs, alcohol and fruit juice, ml/d | 335 (277) | 1 | 14 | 36 | 119 | 278 | 480 | 705 | 864 | 1,226 |
| Non-whole grain foods, RA/d | 1.7 (0.8) | 0.2 | 0.5 | 0.7 | 1.1 | 1.6 | 2.1 | 2.7 | 3.0 | 3.8 |
| Other low nutritive value foods, RA/d | 4.1 (2.7) | 0.7 | 1.1 | 1.5 | 2.2 | 3.4 | 5.2 | 7.5 | 9.2 | 13.4 |
| **Total*^2^*** | | | | | | | | | | |
| Total foods, RA/d | 13.2 (4.1) | 6.1 | 7.6 | 8.5 | 10.3 | 12.7 | 15.5 | 18.6 | 20.7 | 25.5 |
| *^1^*All estimated means and percentiles reflect usual or long-term dietary intakes and were modelled jointly using the National Cancer Institute Markov Chain Monte Carlo (MCMC) multivariate method (see Methods section). Adapted from Statistics Canada, Canadian Community Health Survey- Nutrition: Public Use Microdata File, 2015, June 2023. This does not constitute an endorsement by Statistics Canada of this product. CCHS, Canadian Community Health Survey; CFG, Canada's Food Guide 2019; d, day; HEFI-2019, Healthy Eating Food Index-2019; RA, Reference Amounts (amount of food usually eaten by an individual at one sitting). | | | | | | | | | | |
| *^2^*The total includes all categories of foods and milk, but excludes water and healthy beverages as well as SSBs, alcohol and fruit juice. 1 RA of milk equals to 250 ml. | | | | | | | | | | |

**Supplemental Table 3.** Estimated means and percentiles of nutrients considered in the HEFI-2019 score in adults aged 65 years or more from the CCHS 2015 - Nutrition.*^1^*

|  |  | **Percentile** | | | | | | | | |
| --- | --- | --- | --- | --- | --- | --- | --- | --- | --- | --- |
| **Nutrients** | **Mean (SD)** | 1 | 5 | 10 | 25 | 50 | 75 | 90 | 95 | 99 |
| MUFA, g | 23 (7) | 10 | 13 | 15 | 18 | 22 | 27 | 32 | 36 | 44 |
| PUFA, g | 13 (4) | 6 | 8 | 9 | 10 | 12 | 15 | 18 | 20 | 24 |
| SFA, g | 21 (7) | 9 | 11 | 13 | 16 | 20 | 24 | 30 | 33 | 40 |
| SFA, %E | 11.1 (2.0) | 7.0 | 8.0 | 8.6 | 9.7 | 11.0 | 12.4 | 13.8 | 14.7 | 16.6 |
| Free sugars, g | 46 (26) | 7 | 14 | 18 | 28 | 41 | 58 | 79 | 95 | 132 |
| Free sugars, %E | 10.7 (4.5) | 2.2 | 4.1 | 5.3 | 7.5 | 10.3 | 13.4 | 16.6 | 18.7 | 23.2 |
| Sodium, mg | 2,431 (621) | 1,277 | 1,530 | 1,684 | 1,977 | 2,367 | 2,824 | 3,269 | 3,551 | 4,095 |
| Sodium, g/1000 kcal | 1.5 (0.2) | 1.0 | 1.1 | 1.2 | 1.3 | 1.5 | 1.6 | 1.8 | 1.9 | 2.2 |
| Energy, kcal | 1,663 (428) | 862 | 1,043 | 1,152 | 1,355 | 1,618 | 1,926 | 2,239 | 2,440 | 2,833 |
| *^1^*All estimated means and percentiles reflect usual or long-term dietary intakes and were modelled jointly using the National Cancer Institute Markov Chain Monte Carlo (MCMC) multivariate method (see Methods section). Adapted from Statistics Canada, Canadian Community Health Survey- Nutrition: Public Use Microdata File, 2015, June 2023. This does not constitute an endorsement by Statistics Canada of this product. CCHS, Canadian Community Health Survey; d, day; E, energy; HEFI-2019, Healthy Eating Food Index-2019; %E, percent of total energy. | | | | | | | | | | |

#### Relationship between food and beverage categories of the HEFI-2019 and nutrient intakes

**Supplemental Figure 1** presents the energy-adjusted correlation among food and beverage categories contributing to the HEFI-2019 and nutrient intakes.

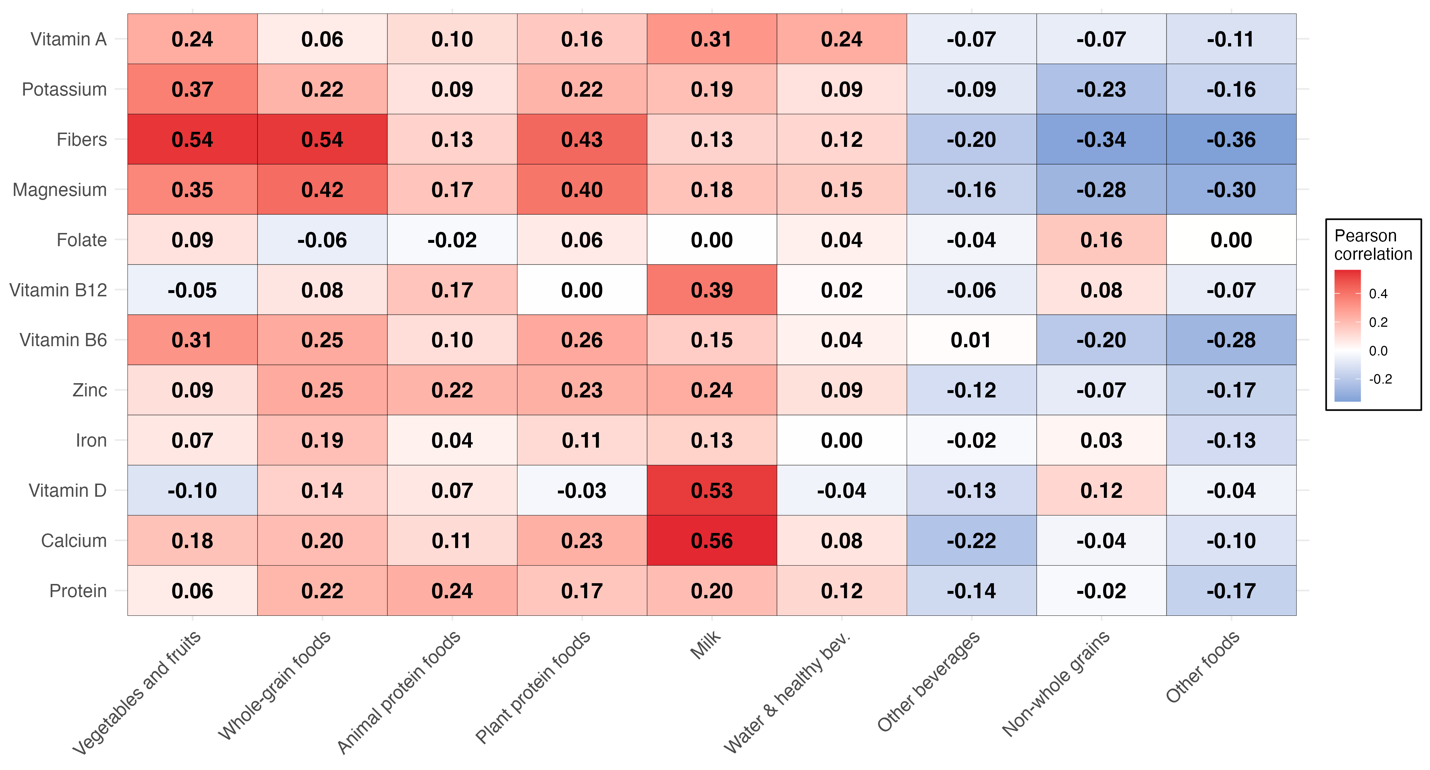

***Supplemental Figure 1:*** *Pearson correlations heat map among food and beverage categories contributing to the HEFI-2019 and nutrient intakes in adults aged 65 years or more from the CCHS 2015 - Nutrition. A higher correlation indicates that higher food or beverage intakes are associated with higher nutrient intakes and vice versa. All dietary intakes were modelled using the National Cancer Institute Markov Chain Monte Carlo multivariate method to estimate usual intakes (see Methods). Estimated correlations were all adjusted for total energy intake. Adapted from Statistics Canada, Canadian Community Health Survey- Nutrition: Public Use Microdata File, 2015, June 2023. This does not constitute an endorsement by Statistics Canada of this product. Bev, beverages; CCHS, Canadian Community Health Survey.*

#### Prevalence of inadequate nutrient intake

**Supplemental Table 4.** Cut-offs (EAR) used to assess the prevalence of intake inadequacy and prevalence estimates (Pr(X<EAR)) in adults aged 65 years or more from the CCHS 2015 - Nutrition*^1^*

|  | **Dietary Reference Intake (DRI) age and sex group** | | | |
| --- | --- | --- | --- | --- |
| **Nutrient** | Males, 65-70 y | Males, 71 y+ | Females, 65-70 y | Females, 71 y+ |
| **Protein** | | | | |
| EAR, g/kg | 0.66 | 0.66 | 0.66 | 0.66 |
| Pr(X<EAR), % | 3.6 (0.0, 8.6) | 9.7 (4.7, 14.6) | 11.5 (4.2, 18.9) | 14.6 (8.9, 20.4) |
| **Calcium** | | | | |
| EAR, mg | 800 | 1000 | 1000 | 1000 |
| Pr(X<EAR), % | 55.0 (46.2, 63.7) | 86.9 (82.0, 91.8) | 87.5 (82.6, 92.4) | 93.9 (90.5, 97.2) |
| **Vitamin D** | | | | |
| EAR, mcg | 10 | 10 | 10 | 10 |
| Pr(X<EAR), % | 89.4 (85.0, 93.9) | 95.6 (92.3, 99.0) | 99.3 (98.1, 100.5) | 99.4 (97.9, 100.8) |
| **Iron** | | | | |
| EAR, mg | 6 | 6 | 5 | 5 |
| Pr(X<EAR), % | 0.0 (0.0, 0.3) | 0.0 (0.0, 0.5) | 0.7 (0.2, 1.1) | 1.1 (0.2, 2.0) |
| **Zinc** | | | | |
| EAR, mg | 9.4 | 9.4 | 6.8 | 6.8 |
| Pr(X<EAR), % | 13.0 (0.4, 25.7) | 44.0 (34.7, 53.4) | 18.1 (9.3, 26.9) | 33.4 (24.4, 42.4) |
| **Vitamin B6** | | | | |
| EAR, mg | 1.4 | 1.4 | 1.3 | 1.3 |
| Pr(X<EAR), % | 15.7 (6.5, 25.0) | 33.0 (24.0, 42.1) | 36.4 (27.6, 45.3) | 58.0 (50.5, 65.6) |
| **Vitamin B12** | | | | |
| EAR, mcg | 2 | 2 | 2 | 2 |
| Pr(X<EAR), % | 0.0 (0.0, 1.1) | 0.5 (0.0, 2.8) | 7.9 (0.0, 17.9) | 11.5 (0.9, 22.2) |
| **Folate** | | | | |
| EAR, mcg | 320 | 320 | 320 | 320 |
| Pr(X<EAR), % | 8.0 (0.0, 16.3) | 14.3 (6.0, 22.6) | 39.7 (28.0, 51.5) | 48.6 (38.1, 59.1) |
| **Magnesium** | | | | |
| EAR, mg | 350 | 350 | 265 | 265 |
| Pr(X<EAR), % | 63.9 (55.1, 72.7) | 80.4 (74.9, 85.8) | 44.7 (35.9, 53.6) | 64.8 (58.8, 70.7) |
| **Fibers*^2^*** | | | | |
| AI, g | 30 | 30 | 21 | 21 |
| **Potassium*^2^*** | | | | |
| AI, mg | 3400 | 3400 | 2600 | 2600 |
| **Vitamin A** | | | | |
| EAR, RAE | 625 | 625 | 500 | 500 |
| Pr(X<EAR), % | 37.1 (19.9, 54.3) | 44.3 (31.2, 57.4) | 24.6 (14.4, 34.9) | 36.9 (26.8, 47.1) |
| *^1^*All nutrients were modelled using the National Cancer Institute Markov Chain Monte Carlo multivariate method to estimate usual intakes (see Methods). Adapted from Statistics Canada, Canadian Community Health Survey- Nutrition: Public Use Microdata File, 2015, June 2023. This does not constitute an endorsement by Statistics Canada of this product. AI, adequate intakes; CCHS, Canadian Community Health Survey; DRI, Dietary Reference Intake; EAR, Estimated Average Requirements; RAE, Retinol Activity Equivalent; X, usual nutrient intake. | | | | |
| *^2^*Prevalence of inadequate intakes are not calculated for potassium and fibers which do not have Estimated Average Requirements due to insufficient evidence, but only Adequate Intakes (AI) values. | | | | |

#### Relationship between the HEFI-2019 score and prevalence of inadequate nutrient intake

##### Prevalence of inadequate protein intake for hypothetical higher recommendations

The current Estimated Average Requirements (EAR) for protein is 0.66 g/kg, based on a Recommended Daily Allowance (RDA) of 0.8 g/kg to avoid negative nitrogen balance. Emerging consensus support that the RDA should be set to at least 1.0 to 1.2 g/kg for healthy older adults to prevent sarcopenia (Bauer et al. 2013; Phillips, Chevalier, and Leidy 2016; Traylor, Gorissen, and Phillips 2018). Accordingly, hypothetical EAR cut-offs for these higher RDA could be approximately 0.8 g/kg (RDA of 1.0 g/kg) or 1.0 g/kg (RDA of 1.2 g/kg).

**Supplemental Figure** **2** presents the relationship between the HEFI-2019 and protein intake inadequacy for the current and hypothetical EAR. HEFI-2019 score at the 90^th^ percentile compared with median HEFI-2019 score reduced the prevalence of inadequate protein intakes for all cut-offs

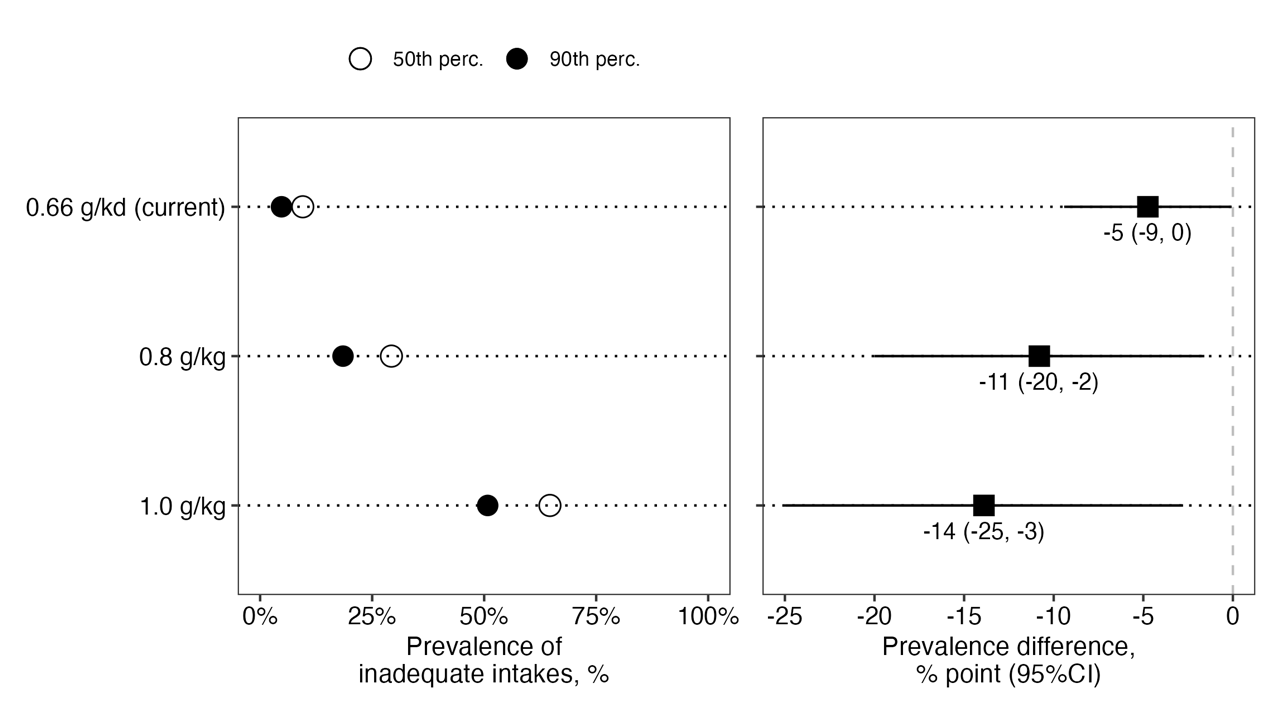

**Supplemental Figure 2:** Prevalence of inadequate protein intakes and difference for HEFI-2019 scores at the 90th compared with the 50th percentile of the score distribution in adults aged 65 y or more from the CCHS 2015 - Nutrition. Inadequate intakes are intakes below the cut-offs (i.e., 0.66, 0.8 and 1.0 g/kg). Both the HEFI-2019 score and nutrient intakes were modeled using the National Cancer Institute multivariate method to assess usual intakes (see Methods). CCHS, Canadian Community Health Survey; HEFI-2019, Healthy Eating Food Index-2019; Perc, percentile.

### Supplementary References

Bauer, Jürgen, Gianni Biolo, Tommy Cederholm, Matteo Cesari, Alfonso J. Cruz-Jentoft, John E. Morley, Stuart Phillips, et al. 2013. “Evidence-Based Recommendations for Optimal Dietary Protein Intake in Older People: A Position Paper From the PROT-AGE Study Group.” *Journal of the American Medical Directors Association* 14 (8): 542–59. <https://doi.org/10.1016/j.jamda.2013.05.021>.

Brassard, Didier, Lisa-Anne Elvidge Munene, Sylvie St-Pierre, Patricia M. Guenther, Sharon I. Kirkpatrick, Joyce Slater, Simone Lemieux, et al. 2022. “Development of the Healthy Eating Food Index (HEFI)-2019 Measuring Adherence to Canada’s Food Guide 2019 Recommendations on Healthy Food Choices.” *Applied Physiology, Nutrition, and Metabolism* 47 (5): 595–610. <https://doi.org/10.1139/apnm-2021-0415>.

Otten, Jennifer J., Jennifer Pitzi Hellwig, and Linda D. Meyers. 2006. *Dietary Reference Intakes : The Essential Guide to Nutrient Requirements*. Book. Washington, D.C.: National Academies Press. <http://www.nap.edu/catalog/11537.html>.

Phillips, Stuart M., Stéphanie Chevalier, and Heather J. Leidy. 2016. “Protein “Requirements” Beyond the RDA: Implications for Optimizing Health.” *Applied Physiology, Nutrition, and Metabolism* 41 (5): 565–72. <https://doi.org/10.1139/apnm-2015-0550>.

Traylor, Daniel A, Stefan H M Gorissen, and Stuart M Phillips. 2018. “Perspective: Protein Requirements and Optimal Intakes in Aging: Are We Ready to Recommend More Than the Recommended Daily Allowance?” *Advances in Nutrition* 9 (3): 171–82. <https://doi.org/10.1093/advances/nmy003>.
